# Diagnostic Accuracy of Fine Needle Aspiration Cytology in Thyroid Diseases: Comparison with Histopathological Findings in Hawassa, Ethiopia

**DOI:** 10.64898/2026.10.16.26365726

**Authors:** Teketel Tadesse Geremew, Abraham Kassahun Tadele, Bizunesh Dires Fenta, Ghion Getenet Engida, Fekade Yerakly Lucas, Tigist Gutema Tesgera, Alemwosen Teklehaimanot Alem, Abebe Melis Nisro

## Abstract

**Introduction:** Thyroid diseases are major health problems and among the most common endocrine disorders worldwide, including in Ethiopia. They are characterized by alterations in hormone secretion, enlargement of the thyroid gland, or both. Fine needle aspiration cytology is an important initial screening tool, and histopathologic examination is essential for the definitive diagnosis of thyroid disease. This study aimed to compare the diagnostic accuracy of fine needle aspiration cytology with histopathology in identifying thyroid diseases.

**Methods:** A cross-sectional study was conducted at three health facilities in Hawassa City, Ethiopia, involving 304 patients who underwent both thyroid fine needle aspiration cytology and histopathologic examination of the same site between 30^th^ October 2024 and 29^th^ October 2025. FNAC results were classified using the Bethesda system, with histopathology as the gold standard. Data were entered into Kobo Toolbox, exported to SPSS version 27, and analyzed using cross-tabulation and chi-square (χ^2^) tests to determine diagnostic accuracy.

**Result:** 304 patient records were included in the study. The fine needle aspiration cytology was determinate/diagnostic in 81.6% of cases. The study revealed the sensitivity and specificity of fine needle aspiration cytology to be 88.2% (CI 77.6-95.1%) and 94.4% (CI 90.6-97.1%), respectively. The false negative and false positive rates in this study were 2.4% and 4.4%, respectively. The positive predictive value was 80.3% (CI 68.7-89.3%), and the negative predictive value was 96.8% (CI 93.8-98.7%). The accuracy of fine needle aspiration cytology in differentiating benign from malignant thyroid lesions was 93.1%.

**Conclusion:** FNAC demonstrated strong agreement with histopathology and remains a reliable, cost-effective first-line diagnostic tool for thyroid disease, particularly in resource-limited settings. Strengthening sample handling, ultrasound guidance, and adherence to Bethesda criteria may further improve diagnostic precision

## Introduction

The normal adult thyroid gland is composed of two lobes joined by the isthmus, which lies across the trachea anteriorly, below the level of the cricoid cartilage^1,2^. The thyroid gland, along with the esophagus, pharynx, and trachea, is found within the visceral compartment of the neck, which is bound by pre-tracheal fascia^2^ . The thyroid gland weighs 10 to 20 grams in normal adults^3^. Thyroid volume measured by ultrasonography is slightly greater in men than women, increases with age and body weight, and decreases with increasing iodine intake ^4^.

The gland is affected in many physiologic and pathologic conditions. Diseases of the thyroid include conditions associated with excessive release of thyroid hormones (hyperthyroidism), thyroid hormone deficiency (hypothyroidism), and mass lesions of the thyroid^5^.

Thyroid nodules are one of the commonest clinical findings, with a reported prevalence of 4-7% in the adult population^6^. Selecting those neoplastic nodules among the different lesions is of paramount importance when deciding to operate on the enlarged thyroid^7^. The incidence varies in different regions depending upon different factors including age, sex, diet, environmental factors, and radiation exposure ^8,9^.

Thyroid disorders are four times more in females than in males. They are endemic in mountainous regions, where the soil, water, and food contain little iodine^10^. Neoplastic diseases are also seen in the thyroid. Adenoma is the commonest benign tumor of the thyroid. Thyroid cancer is the most frequent endocrine malignancy ^11^. Papillary carcinoma is the most common thyroid cancer, followed by follicular, medullary, anaplastic carcinoma, and lymphoma ^12^.

To diagnose different kinds of thyroid disease, a complete clinical examination in addition to hormonal assays and morphology study should be done. In the end, Histopathologic examination provides a definitive diagnosis^13^. One of the well-established and preferred first-line diagnostic tools in the evaluation of diffuse thyroid lesions, as well as of thyroid nodules, is fine needle aspiration cytology (FNAC), with the main purpose of confirming benign lesions and thereby reducing unnecessary surgery ^12^. It is indicated in all palpable thyroid nodules and non-palpable lesions found suspicious on radiology. Non-palpable thyroid nodules are aspirated with ultrasound assistance. Though ultrasound-guided FNAC is relatively expensive, it helps in targeting the area of interest, especially in cystic lesions. ^14^.

FNAC is a minimally invasive, accurate, and cost-effective diagnostic tool for differentiating benign from malignant thyroid lesions, thus providing a basis for decisions on management options. Whereas FNAC has a high degree of proven sensitivity (90%) and specificity (96%), it’s however, associated with a false positive and negative interpretation that averages from 1 to 6% respectively ^12,15^. Papanicolaou International Cytopathology Society guidelines state that acceptable false negative and positive rates should be less than 2% and 3%, respectively ^16^. False negative results in thyroid cancer have the potential to increase patient morbidity due to the delay in diagnosis, and false positive results also may lead to overtreatment of patients, including total thyroidectomy, and sometimes to lawsuits for misdiagnosis and malpractice^17^.

Despite several studies showing a high accuracy of FNAC, studies especially in Eastern Africa and other developing countries with a high prevalence of nodular thyroid disease, have shown the accuracy of FNAC to be lower, and its diagnostic performance has been shown to vary across different studies^18^.

The Bethesda System for Reporting Thyroid Cytopathology, updated in 2017, provides a standardized framework for interpreting FNAC results and guiding patient management^17^. Within this system, nondiagnostic or unsatisfactory samples (Category I) require repeat FNAC with ultrasound guidance, while benign lesions (Category II) are managed with clinical and sonographic follow-up. Cases with atypia of undetermined significance or follicular lesions of undetermined significance (Category III) are recommended for repeat aspiration, molecular testing, or lobectomy. Follicular neoplasms or suspicious follicular lesions (Category IV) are generally managed with molecular testing or lobectomy. Samples suspicious for malignancy (Category V) and malignant lesions (Category VI) are typically treated with near-total thyroidectomy or lobectomy. This classification system ensures consistency in reporting and provides clear clinical pathways for patient care.

This study assessed the diagnostic accuracy of fine needle aspiration cytology to histopathology in thyroid disease among thyroid specimens submitted to Hawassa University Comprehensive Specialized Hospital (HUCSH), Yanet Internal Medicine Specialty Center, and Alatyon General Hospital, Pathology Department, from 30^th^ October 2024 to 29^th^ October 2025.

## Methods

### Study area

The study was conducted at government and private health facilities in Hawassa City, which is the capital of Sidama Regional State. The city is located 275 km southeast of Addis Ababa. It is divided in to 8-sub city administration and with estimated population of 350,000; it covers 50 km^2^ and is located 1,700m above sea level. Hawassa University Comprehensive Specialized Hospital (HUCSH) serves as the teaching hospital at Hawassa University College of Medicine and Health Sciences (HUCMHS). It serves more than 18 million people of the Sidama region, Southern Peoples Regional State, and the surrounding Oromia region zones.

HUCMHS offers undergraduate, postgraduate, PhD, and specialty programs in various departments, including Health Officer, Nursing, Pharmacy, Medicine, Optometry, General Public Health, Epidemiology, Reproductive Health, PhD in Public Health, Internal Medicine, Surgery, Gynecology and Obstetrics, Pediatrics, Pathology, and Ophthalmology. Cytology and histopathology services are available at the pathology departments of HUCSH, Yanet Specialized Medical Center, and Alatyon General Hospital.

### Study Design and Period

An institution-based cross-sectional study was conducted using histopathology reports of patients who had pre-operative thyroid FNAC and subsequently underwent thyroidectomy between October 30, 2024, and October 29, 2025.

### Population, Eligibility, Sample Size Determination, and Sampling Technique/Procedure

The study population consisted of all patients who underwent thyroidectomy and had both FNAC and histopathology reports at three health facilities in Hawassa City: HUCSH, Yanet Specialized Medical Center, and Alatyon General Hospital. Eligible participants were those with pre-operative thyroid FNAC and corresponding post-operative biopsy reports. Patients were excluded if their FNAC or biopsy reports lacked a specific diagnosis or if they had two different FNAC diagnoses recorded.

The sample size was determined using Buderer’s formula to estimate sensitivity and specificity at a 95% confidence level and 5% margin of error. Based on an estimated sensitivity of 96% and prevalence of 39%, the required sample size was 167 (including a 10% non-response rate). Based on an estimated specificity of 92.7% and prevalence of 39%, the required sample size was 294 (including a 10% non-response rate). All eligible patients during the study period were consecutively recruited from the pathology departments of the three institutions using a convenience sampling approach, yielding a total of 304 participants.

### Study variables and Operational definitions

The dependent variable was the diagnostic accuracy of fine needle aspiration cytology in thyroid pathologies. Whereas the independent variables were the size of the thyroid lesion, types of diagnostic procedures (Guided or Conventional) and socio-demographic factors: Age, sex, place of residence **FNAC**: A cytologic diagnostic method based on morphologic findings of individual and small group of cells aspirated using a fine needle by creating a negative pressure.

### Histopathology/biopsy

A procedure by which a tissue sample is taken by a surgical extraction and subsequently processing of sections or whole of the tissue is done to produce thin tissue sections with preserved architecture which can be examined under the microscope after staining.

TP, True positive, FN, False negative, TN, True negative, FP, False positive

### Data collection method, classification and tools

Data was collected using a structured checklist using Kobo Toolbox composed of sociodemographic factors, clinical information, & gross and microscopic information. The sociodemographic factors and clinical information were taken from FNAC and biopsy request formats filled by the treating physicians. The gross and microscopic information was taken from the request format recorded by the pathology residents and seniors. FNAC was done blindly or by ultrasound-guided in the pathology unit. FNAC results were classified according to the 2017 Bethesda system. The Bethesda system was updated in 2017, and patient management recommendations are accordingly summarized in Table 2. Histopathology results were classified based on a classification adopted from previous literature. The discordant cases between FNAC and biopsy were evaluated and resolved by senior pathologists.

**Table 1.** operational definitions for sensitivity, specificity and accuracy of a test.

| Statistical measure | Definition | Calculation |
| --- | --- | --- |
| Sensitivity | ability of a test to correctly identify those with disease | $TP/(TP + FN)$ |
| Specificity | ability of a test to correctly identify those without a disease | $TN/(TN + FP)$ |
| Positive predictive value | the probability that subjects with a positive screening test truly have the disease | $TP/(TP + FP)$ |
| Negative predictive value | the probability that subjects with a negative screening test truly don't have the disease | $TN/(TN + FN)$ |
| Diagnostic accuracy | ability of a test to discriminate between and/or predict disease and health | $TP+TN/(TP+TN+FP+FN)$ |

TP, True positive, FN, False negative, TN, True negative, FP, False positive

**Table 2.** Socio-demographic characteristics of the study participants.

| Variables(N=304) | Category | Frequency | Percent |
| --- | --- | --- | --- |
| Sex | Female | 254 | 83.6 |
|  | Male | 50 | 16.4 |
| Age | ≤20 | 15 | 4.9 |
|  | 21-40 | 182 | 59.9 |
|  | 41-60 | 83 | 27.3 |
|  | >60 | 24 | 7.9 |
| Residence | Urban | 189 | 62.2 |
|  | Rural | 115 | 37.8 |

### Data processing, management and analysis

The cleaned data was exported into Statistical Package for Social Science (SPSS) version 27.0. The study variables were represented in frequency tables and a descriptive summary. The diagnostic value of FNAC was determined by calculating its sensitivity, specificity, positive and negative predictive values, and accuracy. Sensitivity, specificity, predictive value analyses, and accuracy were done with the aid of a 2x2 table. The histopathology results were taken as the gold standard, and the FNAC results that matched the histopathology findings were regarded as true positives. FNAC results with a follicular neoplasm/suspicious for a follicular neoplasm (FN/SFN) were included in the malignant group. Atypia of undetermined significance and suspicious for malignancy were included in the indeterminate group. Descriptive statistics of the data were calculated as percentages and numbers. Kappa coefficient (p=0.047). was done to assess the agreement of FNAC and histopathology.

### Data quality control

Investigators of the study controlled the overall activity. Training was given for the data collection team on the data collection process. Data quality ascertained by designing the proper data collection materials and through continuous supervision. Completeness, accuracy, and consistency of data collection checked on each day of the data collection period. All completed data collection form then examined for completeness and consistency during data entry and analysis.

### Ethical consideration

Ethical clearance and approval of the study was obtained from the institutional review board (IRB) of Hawassa University College of Medicine and Health Science with reference number IRB/372/16. A formal letter was obtained from HUCSH’s chief clinical director and the Department of Pathology to get patients’ medical record charts and collect report formats. The confidentiality of all patient records was maintained. This research involved only retrospective review of medical records without direct patient contact; therefore, adherence to international norms such as the Declaration of Helsinki was not applicable.

## Result

### Demographic Characteristics of Study Participants

The study included 304 patients, of whom 254 (83.6%) were female, with ages ranging from 9 to 80 years (mean 38.6 ± 13.3), most commonly between 21 and 40 years (59.9%), and the majority, 189 (62.2%), residing in urban areas (Table 2).

### Clinical presentation of study participants

Among the 304 patients, the vast majority, 294 (96.7%), presented with an anterior neck mass either alone or in combination with other symptoms, while 66 (21.9%) reported mass-effect symptoms and 54 (17.9%) had features of hyperthyroidism (Table 3). A history of thyroid lesions lasting 1–5 years was noted in 218 patients (71.7%). FNAC was performed for all cases, with 34 procedures guided by ultrasound. Total thyroidectomy was the predominant surgical procedure in 193 cases (63.5%). On gross examination, thyroidectomy specimens ranged from 2 to 18 cm, with a mean size of 7.4 ± 3.8 cm.

**Table 3.** Clinical presentations of the study participants.

| Clinical presentations | Frequency | Percentage |
| --- | --- | --- |
| Anterior neck mass only | 160 | 52.6 |
| Anterior neck mass and hyperthyroidism symptoms | 54 | 17.9 |
| Anterior neck mass and hypothyroidism symptoms | 11 | 3.1 |
| Anterior neck mass and mass effect | 66 | 21.9 |
| Hyperthyroidism | 2 | 0.7 |
| Hypothyroidism | 1 | 0.3 |
| Hypothyroidism, anterior neck mass, and mass effect | 1 | 0.3 |
| Other and anterior neck mass | 2 | 0.6 |
| Other | 7 | 2.6 |
| Total | 304 | 100 |

### Histopathologic and cytopathologic patterns of thyroid lesions

On cytopathologic pattern, most of the thyroid lesions were reported as benign, 192 (63.2%), followed by a report of malignant, constituting 39 (12.5%) cases (Table 4). On histopathology, non-neoplastic lesions were encountered in 202 (66.45%) cases in which nodular colloid goiter (NCG) was the predominant finding. Among the nonneoplastic cases, 176 (87.1%) were NCG, and the remaining 15 (7.4%) cases were thyroiditis, and 11 (5.4%) cases of thyroglossal duct cysts.

**Table 4.** Cytopathologic patterns of thyroid lesions among study participants.

| FNAC diagnosis | Frequency | Percentage |
| --- | --- | --- |
| Benign | 192 | 63.2 |
| Atypia of undetermined significance | 24 | 7.9 |
| Follicular neoplasm | 17 | 5.6 |
| Suspicious for malignancy | 32 | 10.5 |
| Malignant | 39 | 12.8 |
| Total | 304 | 100 |

Neoplastic lesions were encountered in 102 (33.5%) cases, in which benign neoplastic conditions account for 43 (42.1%) cases and malignant neoplastic conditions account for 59 (57.3%) cases.

Follicular adenoma is the most common type among benign thyroid lesions, accounting for 68.1% of benign thyroid lesions, and was reported in 30 patients. Papillary thyroid carcinoma is the most common type among malignant thyroid lesions, reported in 49 patients, and accounts for 83% of malignant thyroid lesions, followed by follicular thyroid carcinoma, reported in 4 patients (6.7%).

### Comparison of FNAC and Histopathologic Examination

Of 24 cases classified as atypia of undetermined significance, 13 were non-neoplastic, 8 benign, and 3 malignant. Among 33 cases suspicious for malignancy, 17 were confirmed as carcinoma, 4 were benign, and 11 were non-neoplastic. Overall, 248 aspirations were diagnostic; 231 (93.2%) howed concordance with histopathology (186 true negatives, 45 true positives), while 17 (6.9%) were discrepant (6 false negatives, 11 false positives). False negatives were all papillary thyroid arcinomas misclassified as NCG. Of the false positives, 2 were follicular-patterned lesions (histologically NCG and congenital anomaly), while 9 cytologically malignant cases proved to be colloid goiter (8) or follicular adenoma (1). FNAC showed a sensitivity of 88.2% (CI 77.6– 95.1%), specificity of 94.4% (CI 90.6–97.1%), positive predictive value of 80.3% (CI 68.7– 89.3%), and negative predictive value of 96.8% (CI 93.8–98.7%). Cyto-histologic correlation is summarized in Figure 1.

**Figure 1.**
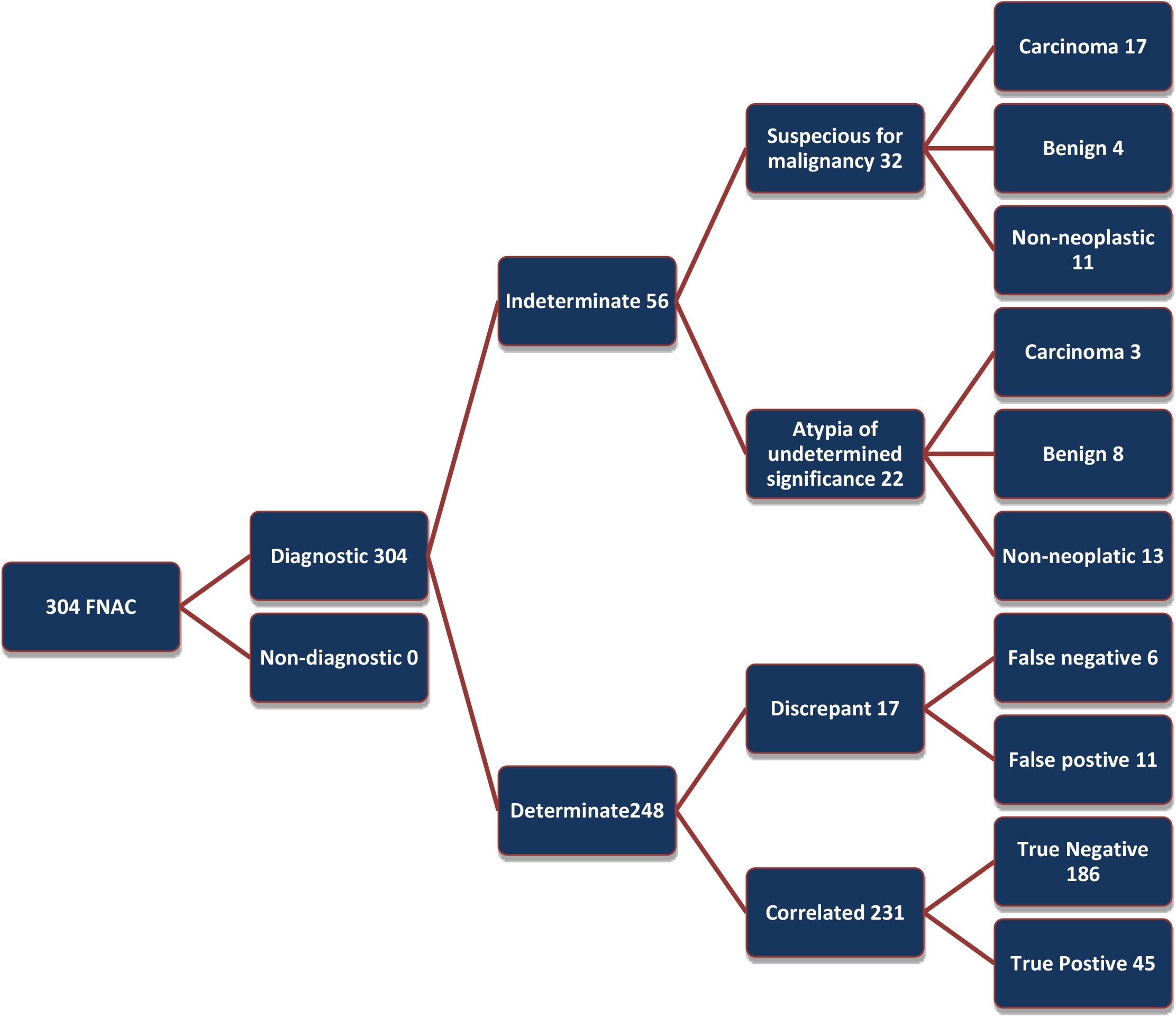
Summary of FNAC results compared with corresponding histopathology findings among 304 study participants.

### Diagnostic accuracy of FNAC against Biopsy on thyroid lesions

The accuracy of FNAC in diagnosing thyroid diseases in this study is 93.1%. FNAC and histopathology showed agreement on diagnosing thyroid diseases at a kappa statistic of 0.798 (Table 5). The diagnostic accuracy of FNAC on thyroid lesions was excellent, with an area under the curve of 0.913 (95% CI,0.859-0.968) at a Youden index of 0.826 (Figure 2).

**Table 5.** Kappa statistics for the diagnostic accuracy of FNAC in thyroid disease.

| Symmetric Measures |  |  |  |  |  |
| --- | --- | --- | --- | --- | --- |
|  |  | Value | Asymptotic Standard Error<sup>a</sup> | Approximate T<sup>b</sup> | Approximate Significance |
| Measure of Agreement | Kappa | 0.798 | 0.047 | 12.582 | .000 |
| N of Valid Cases |  | 248 |  |  |  |
| a. Not assuming the null hypothesis. |  |  |  |  |  |
| b. Using the asymptotic standard error assuming the null hypothesis. |  |  |  |  |  |

**Figure 2.**
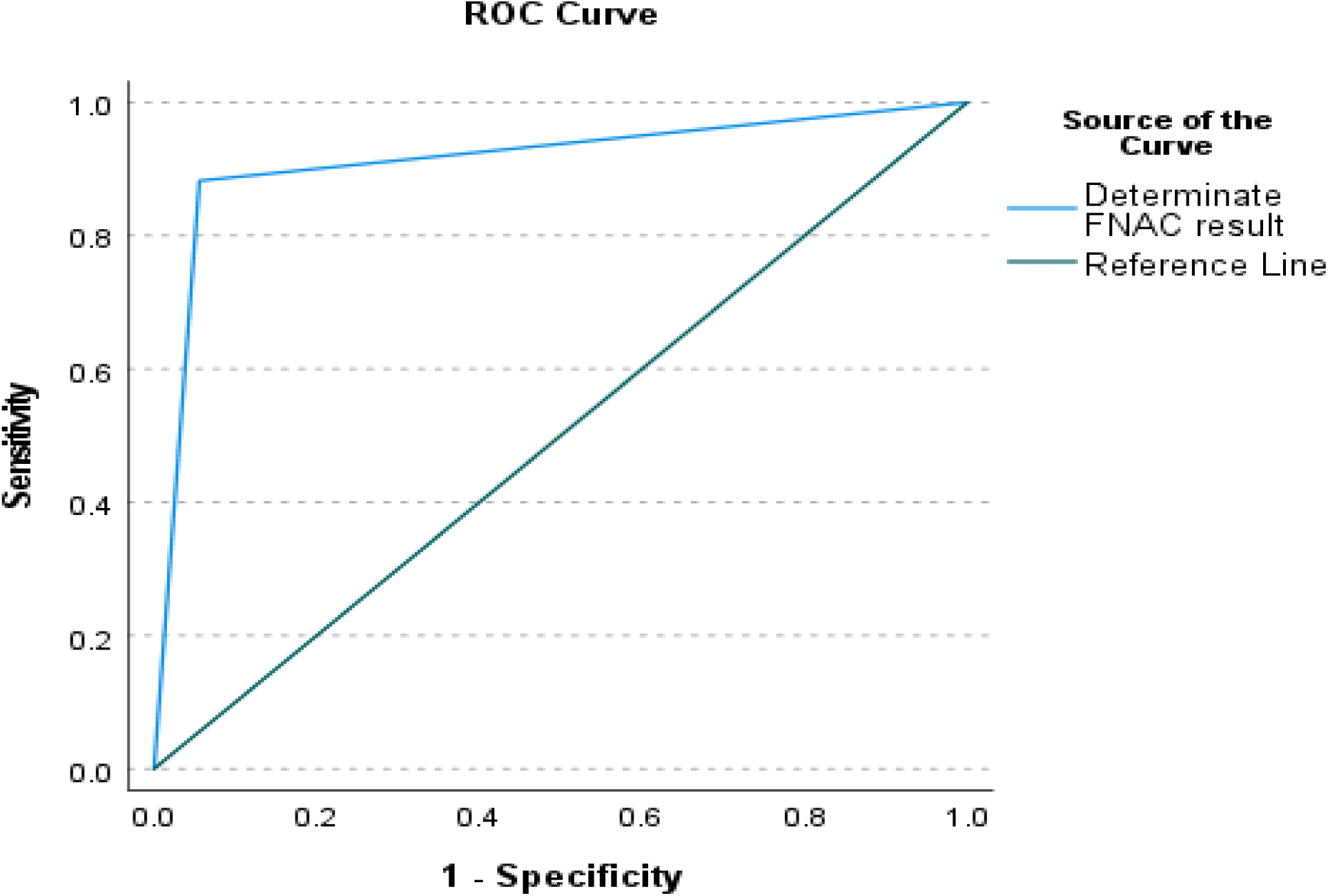
Receiver operating characteristic (ROC) curve for diagnostic accuracy of FNAC.

## Discussion

This study found the diagnostic sensitivity and specificity of FNAC in thyroid lesions to be 88.2% (CI 77.6–95.1%) and 94.4% (CI 90.6–97.1%), respectively. These values are comparable to findings from Jimma, Ethiopia, where sensitivity and specificity were reported as 96% and 92.7% respectively^7^. Findings were also consistent with a cross-sectional analytical study at the Aga Khan University Hospital, Nairobi, Kenya, which reported sensitivity and specificity of 83.2% and 95.2%^19^.

This study’s findings are higher than those from the study at Addis Ababa University, in which the sensitivity and specificity were 67% and 84.7%, respectively^8,20^ the study at the University Teaching Hospital and Ndola Central Hospital in Zambia, in which the sensitivity and specificity were 83.33% and 89.09% ^21^, a study from Lebanon, in which the sensitivity and specificity were 89.31% and 48.44 ^22^, and a study from Philadelphia, USA, in which the sensitivity and specificity were 85.6% and 71.4%^23^.

On the contrary, the finding of this study was lower as compared to the findings of a cross-sectional study done in El-Demerdash Hospital’s surgery department in Egypt, in which the sensitivity and specificity were 100% and 84%, respectively^24^.

These differences in the diagnostic sensitivity and specificity of FNA for thyroid lesions could be because of variation in FNA techniques, use of image/ultrasound-guided FNAC, rapid on-site cytopathologic evaluation, refinements in preparation techniques of cytopathology samples, and improvements to interpretation by the introductions of indeterminate categories, in particular the atypia of undetermined significance/follicular lesion of undetermined significance category, which include a more heterogeneous set of cytological entities by definition, and rates of malignancy exhibit a larger range than originally estimated in the Bethesda document.

False negative and false positive results comprised 6 (2.4%) and 11 (4.44%), respectively. The finding of this study was also found to be comparable to the findings of a similar study done in the USA, in which there were 4% false positives and 8% false negatives.

The finding of this study was found to be lower as compared to the findings in Jimma, Ethiopia, with false negatives and false positives of 22 (8.2%) and 14 (5.2%), respectively, study in an Egyptian study in which the false negative and false positive rates were 7.2% and 5.8%, respectively, study done in Lahore, Pakistan, with false positive rate 35.7% and false negative rate 17.6%^14^.

These differences in the diagnostic false negative and false positive rates of FNA for thyroid lesions could be because of variation in FNA techniques, especially in cystic thyroid lesions, which may lead to a false negative result, the use of image/ultrasound-guided FNAC, rapid on-site cytopathologic evaluation, difference in laboratory set up including staining, drying and availability of equipment’s and improvements to interpretation by strict adherence to adequacy criteria based on the Bethesda system.

The diagnostic accuracy of FNAC in this study was 93.1%, with a negative predictive value (NPV) of 96.8% and a positive predictive value (PPV) of 80.3%. The study finding is comparable with A similar study conducted in Uganda, which showed diagnostic accuracy, NPV, and PPV of 85.9%, 93.9%, and 47%, respectively, ^25^ study done in the USA shows diagnostic accuracy of FNAC to be 86.8% with NPV and PPV of 90% and 85.4%, respectively^23^.

Accuracy is slightly higher than a study done in Jimma and Tikur Anbesa Hospital, Ethiopia, which showed 86.6% and 82% diagnostic accuracy, respectively.^7,20^, study in Lebanon in which the PPV of 78%, NPV of 68.89% ^22^. The study finding was slightly lower as compared to the study done in Diyarbakir, Turkey; accuracy was 93.5%, and positive predictive value and negative predictive value were 91.4% and 93.8%, respectively^26^.

With regard to the risk of malignancy in suspicious or indeterminate groups, 3 (13.6%) out of 22 and 17 out of 32 (53.1) suspicious cases correlated with histopathologic diagnosis of thyroid carcinoma. Thus, there is a significant probability that a cytologically indeterminate lesion could be neoplastic, and there is a good chance that this lesion will be malignant. This is in agreement with a study done in Jimma, Tikur Anbesa Hospital, showing seven out of fifteen suspicious cases (46.7%) and four out of seven suspicious cases (57%) turning out to be malignant, respectively,^7,20^ and a study done in the USA in which 19 out of 40 suspicious cases (47%) were indeed malignant^23^.

### Strengths and Limitations

This multi-facility study with an adequate sample size addresses an important but under-researched topic in the region. However, as a cross-sectional hospital-based design, it was limited by the inability to retrieve cytology slides for reevaluation of discrepant cases and potential selection bias that may reduce generalizability to the wider community.

## Conclusion

This study demonstrates that FNAC is a reliable and effective tool for the initial evaluation of thyroid lesions, showing strong agreement with histopathology and excellent overall diagnostic performance. While occasional misclassification highlights the importance of careful technique and, where possible, ultrasound guidance, the method remains cost-effective and practical in resource-limited settings. By strengthening sample handling and adherence to Bethesda criteria, FNAC can further enhance diagnostic precision and contribute significantly to improved patient care in the region.

## Data Availability

The datasets used or analyzed during the current study are available from the Hawassa University College of Medicine and Health Sciences IRB and corresponding author.

## Availability of data and materials

### Conflict of interest

There is no conflict of interest among authors

### Funding

No funding for this study.

## Notes

### Competing Interest Statement

The authors have declared no competing interest.

### Author Declarations

Ethical clearance and approval of the study was obtained from the institutional review board (IRB) of Hawassa University College of Medicine and Health Science with reference number IRB/372/16. A formal letter was obtained from HUCSH's chief clinical director and the Department of Pathology to get patients' medical record charts and collect report formats. The confidentiality of all patient records was maintained. This research involved only retrospective review of medical records without direct patient contact therefore, adherence to international norms such as the Declaration of Helsinki was not applicable.

